# Music-Induced Cortical Plasticity: Protocol for a Systematic Review

**DOI:** 10.1101/2025.10.21.25338495

**Authors:** Isaiah Osei Duah Junior, Danielle S. Rodriguez, Cassandra Germain

**Author notes:** Correspondence: **Isaiah Osei Duah Junior:** Department of Optometry and Visual Science, College of Science, Kwame Nkrumah University of Science and Technology, Kumasi, Ghana; **Cassandra Germain**: Department of Psychology, College of Health and Human Sciences, North Carolina Agricultural and Technical State University, Greensboro, NC, United States of America.

## Abstract

Music engages sensory, motor, cognitive, and emotional systems, making it a powerful model for studying experience-dependent neuroplasticity. Although research on music-related brain changes is expanding, integration of structural, functional, and cerebrovascular findings remains limited, and effects on higher-order cognitive processes remain unclear. This systematic review will synthesize evidence on music-induced cortical adaptations, including structural changes (e.g., gray and white matter alterations), functional modifications in neural networks, and cerebrovascular dynamics, Associations with behavioral outcomes such as attentional control, executive functioning, language processing and emotional processing across the adult lifespan (18+) will also be examined The protocol follows PRISMA and PRISMA-P guidelines and will be prospectively registered with PROSPERO. Searches will be conducted across PubMed, Scopus, Web of Science, PsycINFO, EMBASE, CENTRAL, and Google Scholar, supplemented by citation tracking of relevant reviews. Eligible studies will include randomized and non-randomized intervention trials and observational designs (longitudinal, case-control, cross-sectional). Primary outcome will be focused on music’s effects on brain structure and activity. Functional and behavioral measures will be analyzed as secondary endpoints. Data extraction and risk of bias assessment will be performed independently by two reviewers using validated instruments (RoB 2.0, ROBINS-I, ROBINS-E, STROBE, and NHLBI tools). A narrative synthesis will be conducted in accordance with SWiM guidelines, with meta-analyses undertaken where appropriate. Certainty of evidence will be assessed using GRADE scale. Collectively, this protocol establishes a rigorous framework to systematically evaluate how music shapes the brain’s structure, function, and vascular systems, and how these changes translate into cognitive and behavioral outcomes.

## Introduction

Music is a unique human activity that engages the brain in a uniquely multimodal manner, simultaneously recruiting sensory, motor, cognitive, and affective systems [1–5]. Beyond its cultural and aesthetic significance, music has emerged as a powerful model for studying neuroplasticity, the brain’s capacity to reorganize its structure, connectivity, and function in response to experience [6–9]. By activating auditory, visual, motor, cognitive, and emotional networks in parallel [1–4, 10–13], music provides a rich, multisensory stimulus for probing experience-dependent neural plasticity across the lifespan [14–18].

Although evidence for the cognitive benefits of music training remains mixed [19–21], converging neural data consistently demonstrate structural and functional cortical adaptations associated with music training and engagement [22–29]. Structurally, music exposure has been linked to increased gray matter volume and cortical thickness in sensory and motor cortices, enhanced white matter integrity in tracts such as the corpus callosum and arcuate fasciculus, and region-specific cortical map reorganization [22–27]. Functionally, music engagement supports enhanced auditory discrimination [28–32], improved sensorimotor integration [33–35], refined executive function [36], and strengthened intra-and inter-network connectivity [37–39]. These adaptations have been observed across the lifespan, from infants exposed to music early in life to older adults in whom music may promote cognitive resilience [22–28, 36, 40, 41].

Behavioral findings complement this neural evidence, with music training linked to superior performance in language acquisition [42–44], verbal memory [42–44], attentional control [45, 46], emotional regulation [47, 48], social communication [49, 50], and motor coordination [51]. Additionally, music exposure has been associated with improved academic achievement [52], enhanced well-being, and mitigation of age-related cognitive decline [53, 54]. Collectively, these findings illustrate how music-induced structural and functional plasticity can yield meaningful real-world benefits, as summarized in **Figure 1**.

**Figure 1:**
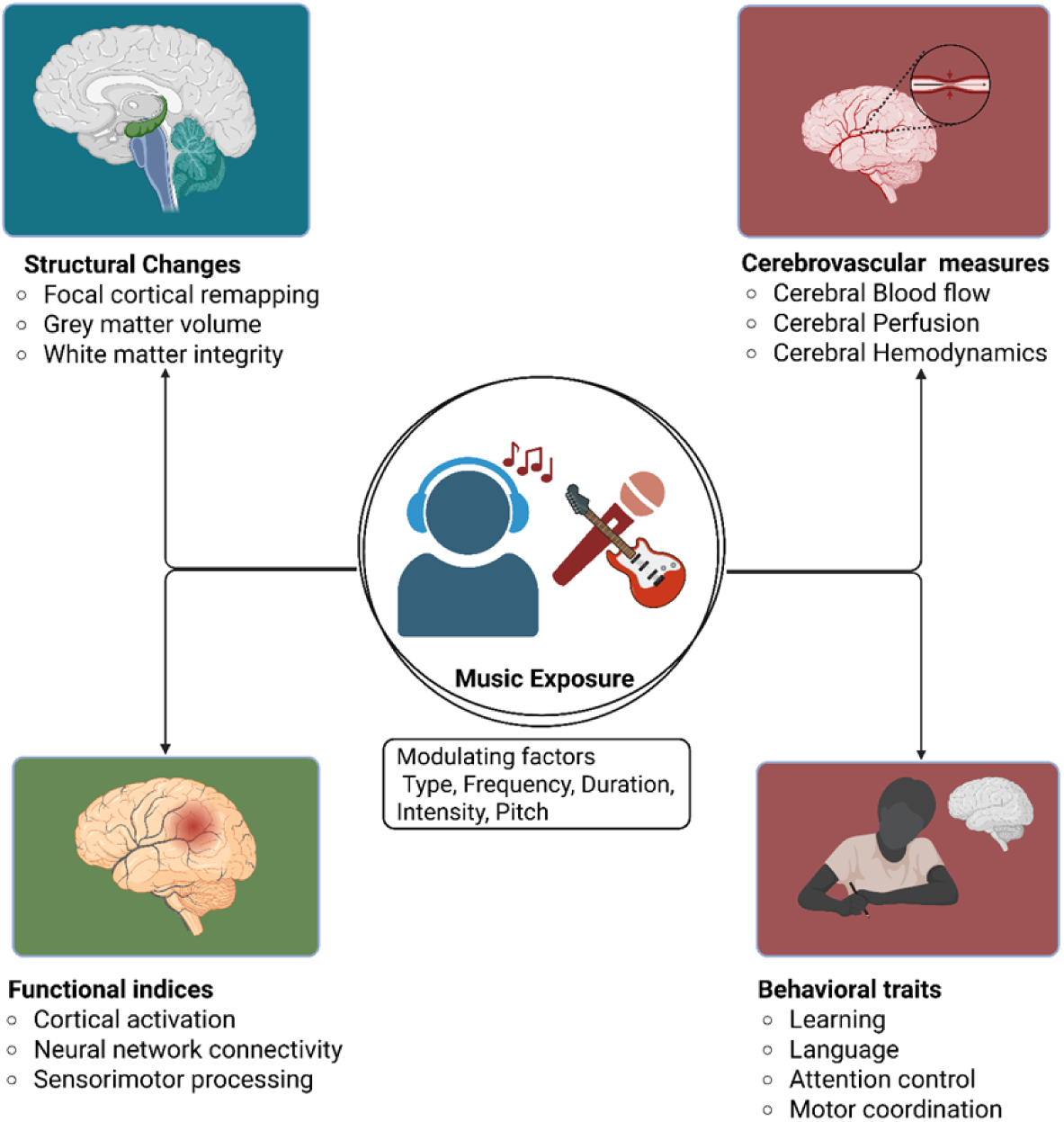
Multidimensional effects of music exposure on the brain and behavior

Despite notable advances, the evidence base remains highly fragmented, largely due to marked heterogeneity in study populations, intervention protocols, and assessment methodologies. Participant samples vary widely, with some investigations targeting adults or children [22, 25, 29, 55, 56], encompassing both musicians and non-musicians [56–61], while others specifically examine the short-and long-term effects of music-based interventions [29, 56, 62, 63]. The modalities of music engagement under study are similarly diverse, ranging from instrumental practice and singing to passive listening and structured music-based therapies [64–66]. Neural outcomes have been characterized using a broad spectrum of techniques, including diffusion tensor imaging (DTI) [67, 68], electroencephalography (EEG) [69–71], magnetoencephalography (MEG) [72–74], and structural and/or functional magnetic resonance imaging (MRI) [25, 64, 75–78]. Together, this methodological and conceptual heterogeneity complicates interpretation, constrains cross-study comparability, and ultimately impedes the formulation of generalizable conclusions regarding the nature, extent, and persistence of music-induced cortical plasticity, as well as its translational potential for therapeutic innovation.

Existing reviews and meta-analyses have yielded valuable insights but remain limited in scope Most have focused primarily on music’s effects within auditory and motor cortices, regions central to perception and production [1, 79]. Yet emerging evidence indicates that music also induces plasticity in visual, prefrontal, parietal, and limbic cortices. These higher-order regions support multisensory integration, executive control, attention, and emotional regulation, functions essential for broad cognitive and affective outcomes. Despite this, no synthesis to date has comprehensively evaluated music’s impact on these domains, leaving a critical gap in the literature. Similarly, prior reviews disproportionate emphasis on behavioral and cognitive outcomes, such as memory [80, 81], attention [82, 83], language [84, 85], and emotional regulation [47, 86]. While informative, these outcomes offer only indirect evidence of the neural mechanisms underlying music’s effects. Without systematic integration of structural and functional findings, the neurobiological basis of music-induced change remains poorly understood. Addressing this gap is essential, as delineating cortical adaptations provides direct mechanistic insight and advances the field beyond descriptive accounts of behavior.

In addition to structural and functional adaptations, emerging evidence suggests that music engagement may influence cerebral blood flow (CBF), a critical physiological marker of cortical activity and plasticity [87–89]. CBF reflects the brain’s metabolic demands and vascular responsiveness [90], providing a dynamic index of localized neural activity that supports synaptic remodeling [91]. By simultaneously engaging sensorimotor networks, music has the potential to modulate regional perfusion, enhance neurovascular coupling, and facilitate experience-dependent cortical reorganization [92–94]. Despite its importance, the relationship between music-induced plasticity and CBF remains poorly understood, leaving the vascular and hemodynamic dimensions understudied. Systematic reviews that integrate CBF metrics with measures of cortical plasticity are particularly promising, as they can consolidate fragmented findings, reveal consistent patterns across diverse study designs, and provide mechanistic insights linking neural remodeling to functional and metabolic dynamics.

Clarifying these neural mechanisms carries both theoretical and practical significance. Mapping how music reshapes cortical architecture and connectivity advances fundamental principles of neuroplasticity while informing translational innovation. If music reliably induces adaptive plasticity in circuits supporting sensory integration, executive control, or emotion regulation, it can be leveraged to develop novel, neuroscience-based interventions. Its cultural ubiquity, motivational appeal, and multimodal reach make music uniquely suited as both a research tool and a clinical strategy for neurological and psychiatric conditions, including stroke, dementia, depression, and developmental disorders.

Collectively, these considerations highlight the need for a systematic review that critically consolidates evidence on music and cortical plasticity. By applying transparent, rigorous, and reproducible methods, such a review can synthesize findings across structural and functional domains, examine effects across auditory, motor, visual, and higher-order associative cortices, and integrate these with emerging mechanistic insights. This approach will clarify inconsistencies, identify gaps for future research, and highlight methodological strengths and limitations. The primary aim of the present review is to evaluate the impact of music engagement on cortical structure, focusing on gray and white matter, cortical thickness, connectivity, and cerebrovascular changes across diverse populations. The secondary aim is to synthesize evidence on the functional and behavioral correlates of these structural adaptations, including their influence on cognition, emotional wellbeing and vision.

## Materials and Methods

This systematic review will be conducted in accordance with the Preferred Reporting Items for Systematic Reviews and Meta-Analyses (PRISMA) guidelines to ensure comprehensive, transparent, and standardized reporting [95]. In instances where quantitative synthesis or meta-analysis is not feasible due to heterogeneity in study designs, populations, interventions, or outcomes, the Synthesis Without Meta-analysis (SWiM) reporting framework will be applied to guide the narrative synthesis and to enhance the clarity, transparency, and reproducibility of the evidence synthesis process [96]. The review protocol will further adhere to the Preferred Reporting Items for Systematic Review and Meta-Analysis Protocols (PRISMA-P) checklist (**S1 Table**) to provide a structured framework for the design, planning, and prospective reporting of methods [97, 98]. In addition, the protocol is prospectively registered with the International Prospective Register of Systematic Reviews (PROSPERO: CRD420251159362) to ensure methodological rigor, prevent unnecessary duplication, and promote accountability and transparency in the review process.

### Eligibility Criteria

#### Inclusion criteria

The eligibility criteria for this systematic review will be defined according to the PICOS framework, encompassing the population, intervention, comparators, outcomes, and study design. **Population (P)**

This review will include studies enrolling human participants across the lifespan, specifically adolescents, adults, and older individuals, while excluding infants and young children due to rapid, dynamic brain development that may confound interpretation of music-induced adaptations. Both healthy and clinical populations (e.g., individuals with neurological, psychiatric, or developmental conditions) will be eligible, provided music exposure or training is part of the intervention. The review will consider both musicians and non-musicians. Musicians are defined as individuals with documented formal or informal training, practice, or sustained engagement in music performance or study, irrespective of instrument or genre. Non-musicians are defined as individuals without prior formal training who receive music exposure through an intervention. Participants will be included regardless of prior music exposure, sex, gender, ethnicity, socio-economic status, language, cultural background, or geographical location.

#### Intervention/Exposure (I)

Studies that evaluate different forms of music exposure (see **Table 1** for a comprehensive list), including but not limited to instrumental practice, vocal training, structured lessons, ensemble participation, and music listening. Both short-term and long-term exposures will be considered, as well as observational studies examining the extent and type of musical experience.

**Table 1:**
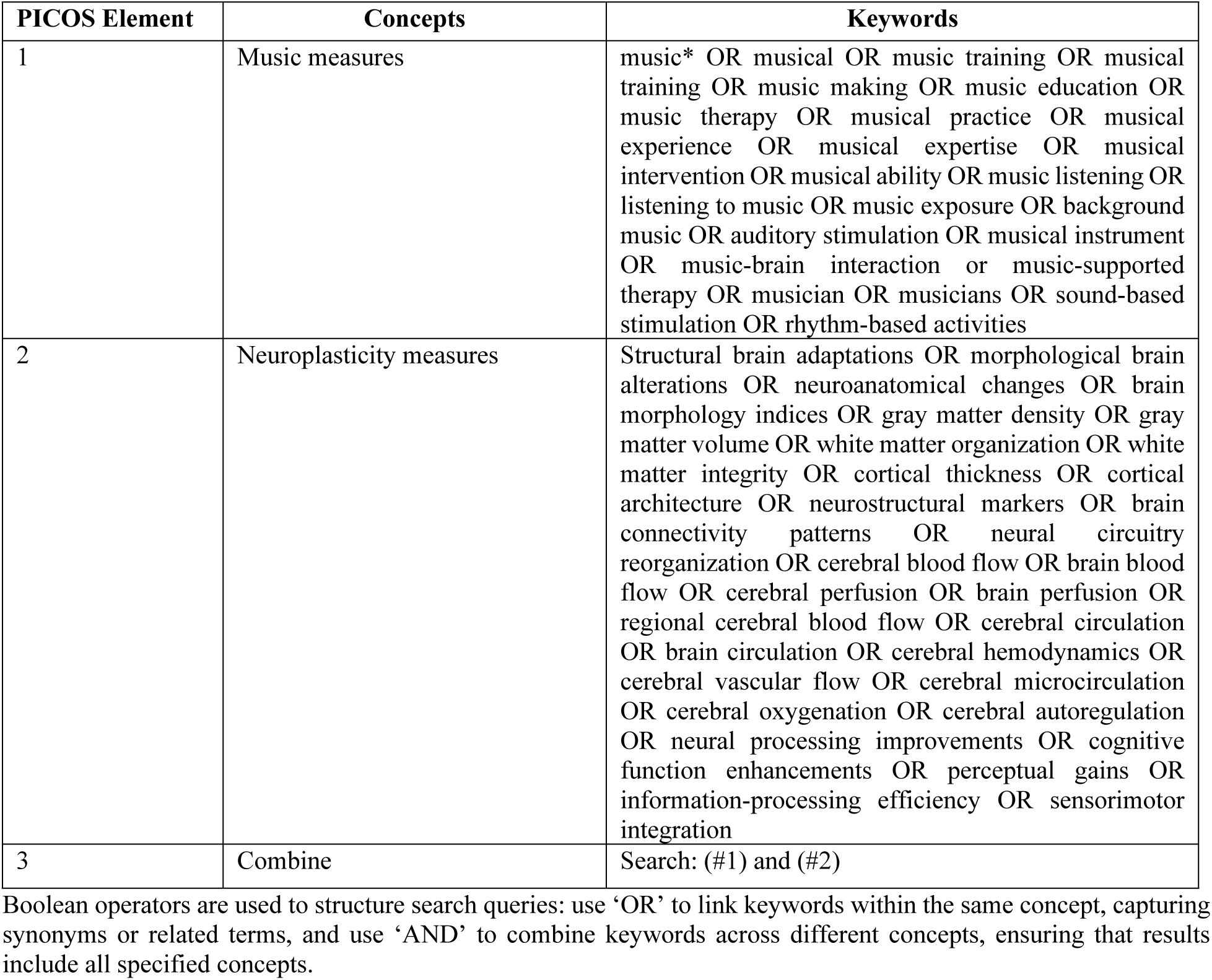
Search strategy developed to be adapted for all databases.

#### Comparison (C)

Comparators for the study where appropriate will be the control arm with that comprises of individuals without prior music training or exposure (music-naïve or untrained participants), participants receiving no intervention, and those engaged in alternative enrichment activities such as sports, visual arts, reading, or other structured programs. These comparison conditions will be included to determine whether the observed effects are specific to music engagement or reflect general benefits of enrichment activities. Studies will be eligible if they employ cross-sectional, longitudinal, randomized, or quasi-experimental designs in which participants with music experience are compared to one or more of these control groups. Alternative activity controls will vary in structure and intensity (e.g., weekly sports practice, art lessons, reading interventions), providing a basis to evaluate the specificity of outcomes to music training.

#### Outcomes (O)

The primary outcome of interest will be structural brain adaptations and/or changes, including gray matter volume and density, white matter integrity, cortical thickness and architecture, brain connectivity patterns, and neural circuitry reorganization. In addition, cerebral blood flow and hemodynamic measures will be included as primary indicators of neurovascular changes associated with music engagement. Secondary outcomes will include functional and behavioral/cognitive changes. The functional changes will encompass improvements in auditory processing, motor coordination, working memory, language, attention-control, executive functions, visual processing, and sensorimotor integration. Behavioral and cognitive performance outcomes will include learning efficiency, skill acquisition, academic achievement, social competence, cognitive-behavioral outcomes, and adaptive functioning.

#### Study Design (S)

The review will include a broad range of study designs to capture the diverse evidence on music-related plasticity and brain outcomes. These will include cross-sectional studies in which musicians are compared to non-musicians or individuals without formal music training to identify potential group differences in cognitive, behavioral, or neural outcomes, thereby allowing the exploration of associations between accumulated musical experience and markers of brain structure or function. The review will also include longitudinal intervention studies, where participants undergo structured music training or music-based activities over a defined period with both pre-and post-intervention assessments, providing insights into causal effects of music engagement on neural plasticity and cognitive or behavioral change. In addition, randomized controlled trials (RCTs), regarded as the gold standard for causal inference, will be incorporated where music-based interventions such as instrumental training, singing, ensemble participation, or music interventions are compared to active or passive control groups such as non-musical enrichment, leisure activities, or no intervention, thereby helping to determine the specificity and robustness of music-related effects. The review will further include neuroimaging studies employing modalities such as functional magnetic resonance imaging (fMRI), electroencephalography (EEG), magnetoencephalography (MEG), diffusion tensor imaging (DTI), or structural MRI when they examine the structural and functional correlates of music engagement. These studies may adopt either cross-sectional or longitudinal designs but must explicitly assess neural outcomes linked to music training or experience. By including this variety of designs, the review will capture both correlational and causal evidence, spanning behavioral and neurobiological levels of analysis.

### Exclusion Criteria

The review will exclude studies if they involve human participants under eighteen years (given the temporal changes in brain development), non-human primates, as this review focuses exclusively on human participants from adolescence to older adults. Research that does not include music engagement or where the effects of music cannot be independently assessed, such as interventions combined with other modalities without isolating music-specific contributions, will also be excluded. Additionally, studies lacking a comparison group or appropriate control conditions will not be considered, as these designs do not allow for the evaluation of causal or relative effects of music engagement. Studies will be excluded if they do not report measurable outcomes related to structural or functional neuroplasticity, cognitive functions, or behavioral and psychosocial domains associated with music engagement; those reporting only subjective experiences or anecdotal observations will similarly be excluded. Finally, publication types such as reviews, commentaries, editorials, conference abstracts without full datasets, methodological or theoretical papers without original empirical findings, and non-peer-reviewed or unpublished studies without sufficient data will be excluded. These criteria ensure that only high-quality, empirically rigorous studies providing meaningful evidence on the effects of music engagement are included.

### Database Sources and Search Strategy

A comprehensive search will be conducted in electronic databases including PubMed, Scopus, Web of Science, PsycINFO, EMBASE, Google Scholar, and the Cochrane Central Register of Controlled Trials (CENTRAL) from inception to date, without restrictions on language or publication year. Additional sources will include reference lists of eligible studies, relevant reviews, and gray literature such as conference proceedings and dissertations. Search terms will be derived from the PICOS framework and will include combinations of keywords and controlled vocabulary (e.g., MeSH terms) related to music exposure, training, or engagement; neuroplasticity, brain structure, brain function, or cognitive outcomes; and human participants. Boolean operators (“AND,” “OR”) will be used to combine terms as shown in **Table 1**.

### Study Selection

The studies retrieved from the databases will be organized by database using the “My Groups” feature in EndNote Reference Manager Version 20 and subsequently exported to Covidence for comprehensive screening based on a priori eligibility criteria. Specifically, two reviewers (I.O.D.J. and D.S.R.) will independently screen all titles and abstracts to determine their relevance to the research question. Studies that appear to meet the inclusion criteria, or those for which the abstract provides insufficient information to make a determination, will be retrieved in full text for further evaluation. Each full-text article will then be assessed independently by the two reviewers against the pre-specified inclusion and exclusion criteria. Any discrepancies or disagreements between the reviewers at either the screening or full-text review stage will be resolved through discussion and consensus. If consensus cannot be reached, a third reviewer (C.G.) will be consulted to make a final decision. The entire study selection process, including the number of records identified, screened, excluded, and included, will be systematically documented using a PRISMA flow diagram to ensure transparency and reproducibility as shown in **Figure 2**.

**Figure 2:**
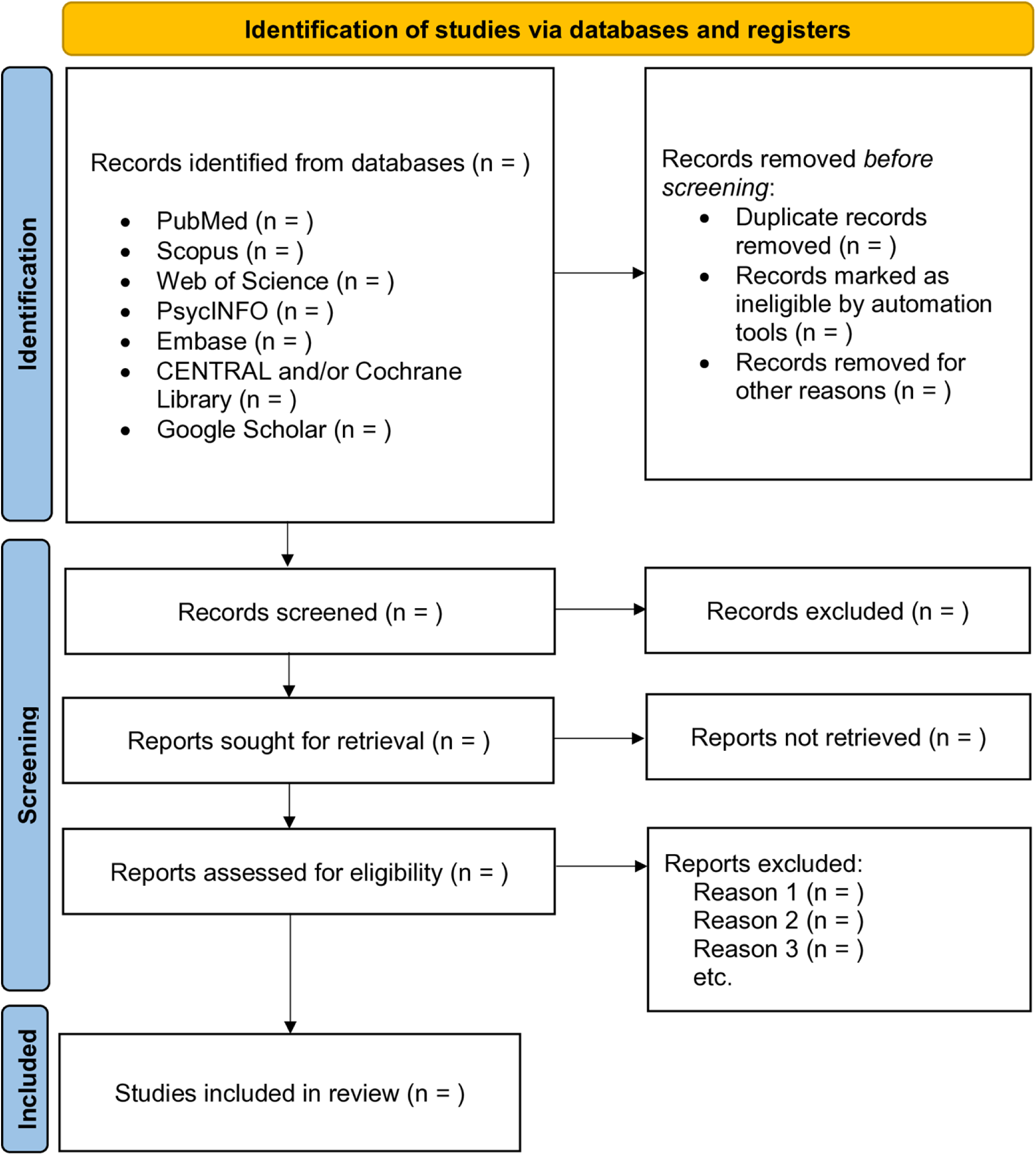
P**R**SIMA **flow diagram for study selection**

### Data Extraction

Data will be independently extracted by two reviewers (I.O.D.J. and C.G.) using a standardized extraction form. The extracted information will include study characteristics such as author, year, country, study design, sample size, age range, and population. Details of the intervention or exposure will be collected, including the type, duration, frequency, and intensity of music engagement or training. Comparison conditions will be recorded, including the type of control group or alternative activity. Outcome measures will encompass neuroplasticity indicators (structural, functional, and cerebrovascular) together with cognitive and behavioral performance metrics, and assessment methods. The key findings and statistical results will be extracted and synthesized. Any discrepancies in data extraction will be resolved through consensus or, if necessary, with input from a third reviewer.

### Risk of Bias

The risk of bias of included studies will be assessed independently by two reviewers (I.O.D.J. and D.S.R.), with any disagreements resolved through discussion or consultation with a third reviewer (C.G.). The assessment tools will be selected according to study design to ensure a rigorous evaluation. For randomized controlled trials (RCTs), the Cochrane Risk of Bias Tool (RoB 2.0)will be used to evaluate potential biases across key domains, including the randomization process, deviations from intended interventions, missing outcome data, outcome measurement, and selection of reported results, with each domain rated as “low risk,” “some concerns,” or “high risk,” and an overall risk-of-bias judgment assigned for each study [99]. For non-randomized interventional studies, the ROBINS-I tool (Risk Of Bias In Non-randomized Studies - of Interventions) will be used to assess bias across domains such as confounding, selection of participants, classification of interventions, deviations from intended interventions, missing data, outcome measurement, and selection of reported results [100]. Similarly, the Risk Of Bias In Non-randomized Studies of Exposures (ROBINS-E) will assess bias in observational studies examining exposure effects across multiple domains, including confounding, participant selection, exposure classification, deviations from intended exposures, missing data, outcome measurement, and selective reporting, rating each as “low” “some concerns,” or “high” risk, with an overall judgment provided for the study [101]. For observational studies, adherence to the Strengthening the Reporting of Observational Studies in Epidemiology (STROBE) checklist will be adopted to guide the evaluation of study quality and reporting transparency [102].

### Quality Assessment

The methodological quality of the studies included in the review will be assessed using the National Heart, Lung, and Blood Institute (NHLBI) quality assessment tools to evaluate overall study (See link: https://www.nhlbi.nih.gov/health-topics/study-quality-assessment-tools) rigor.

These assessments will consider the clarity of research questions, definition and selection of study populations, sample size justification, validity and reliability of exposure and outcome measurements, appropriateness of statistical analyses, adjustment for confounding variables, and transparency in reporting, with studies classified as “good”, “fair”, or “poor” based on the overall evaluation. For neuroimaging studies, an adapted set of quality assessment criteria will be applied to account for the unique methodological considerations of this research. These criteria will focus on sample size adequacy, participant selection and description, imaging modality and acquisition parameters, preprocessing and analysis pipelines, statistical rigor, correction for multiple comparisons, and clarity of reporting, with studies rated according to methodological rigor and susceptibility to bias. Taken together, this structured, design-specific approach will ensure a comprehensive evaluation of study quality, enhancing the reliability of the evidence synthesis and supporting robust interpretation of findings.

### Data Synthesis

A narrative synthesis will be conducted in accordance with the Synthesis Without Meta-analysis (SWiM) guidelines due to the anticipated heterogeneity across included studies in terms of study design, intervention characteristics, and outcome measures [96]. The narrative synthesis will involve a structured and systematic approach to summarizing findings across studies, highlighting patterns, consistencies, and discrepancies in results. Where feasible, effect sizes, such as Cohen’s d or Hedges’ g, will be extracted from primary studies or calculated from reported statistics including means, standard deviations, t-values, or F-values to allow for quantitative comparison across studies. Structural and functional neuroplasticity outcomes will be summarized according to brain region (e.g., prefrontal cortex, motor cortex, auditory regions, hippocampus), modality (e.g., structural MRI, diffusion tensor imaging, fMRI (functional Magnetic Resonance Imaging), EEG (Electroencephalography), ERG (Electroretinography), ERP (Event-Related Potentials, Eye tracking), and measurement method (e.g., voxel-based morphometry, cortical thickness analysis, connectivity metrics, hemodynamic or electrophysiological indices). Behavioral and cognitive outcomes will be categorized by domain to facilitate interpretation, including memory (working memory, episodic memory), executive function (inhibitory control, cognitive flexibility, planning), attention and processing speed, social and emotional skills (emotional regulation, empathy, interpersonal functioning), and learning and academic performance. Where subsets of studies demonstrate sufficient homogeneity in intervention type, outcome measures, and study design, meta-analyses will be conducted to quantitatively synthesize results [103]. Heterogeneity among studies will be assessed using the I² statistic, with thresholds interpreted as low (25%), moderate (50%), and high (75%) heterogeneity, and random-effects models will be applied to account for expected between-study variability. Sensitivity analyses may be performed to explore the influence of study quality, sample characteristics, or intervention parameters on pooled effect sizes. The publication bias will be assessed using funnel plot visualization and statistically tested using Egger’s regression test, with adjustments considered, such as the trim-and-fill method, if asymmetry is detected. All meta-analysis will be performed in R-studio using *metaprop* function from the R**-**meta-package.

### Confidence in Cumulative Evidence

The certainty of evidence for each outcome will be evaluated using the Grading of Recommendations Assessment, Development, and Evaluation (GRADE) framework [104]. This approach assesses the overall body of evidence across five key domains: risk of bias, inconsistency of results, indirectness of evidence, imprecision, and publication bias. Based on these criteria, evidence will be classified as high, moderate, low, or very low, reflecting the confidence in the results [104]. Employing the GRADE approach will ensure that the review not only synthesizes findings but also communicates the reliability of the evidence, which is essential for guiding culturally relevant interventions.

### Handling of Missing Data

For studies with missing, incomplete, or unclear information, attempts will be made to contact corresponding authors to obtain additional data or clarifications. When such data cannot be retrieved, the nature and extent of the missing information will be clearly documented. For quantitative outcomes, the potential impact of missing data on the results will be assessed, and sensitivity analyses will be performed where possible to evaluate how assumptions regarding missing data influence the findings. Studies with substantial or critical missing data that could undermine the validity of outcomes may be rated lower in quality or excluded from meta-analysis, though their results will still contribute to the narrative synthesis. The presence of missing data will also inform the risk of bias and quality assessments using the Cochrane Risk of Bias 2 tool, ROBINS-I, and the STROBE checklist. By systematically addressing missing data, the review aims to reduce potential bias and ensure conclusions are based on the most complete and reliable evidence available.

## Discussion

This review will synthesize current evidence on the effects of music on the adult brain. Music is a uniquely powerful stimulus, engaging auditory, visual, motor, cognitive, and emotional systems simultaneously. Converging research demonstrates that music can induce structural and functional brain changes [38, 105], yet findings remain fragmented, with many studies addressing isolated outcomes such as neural activity [106, 107], cerebral blood flow [87–89], or behavior [108, 109]. What remains unclear is how these neural, vascular, and behavioral changes integrate, particularly in higher-order regions supporting learning, memory, and social interaction.

The primary aim of this review is to evaluate the evidence for music-induced neuroplasticity, encompassing structural adaptations (e.g., gray and white matter changes), functional dynamics (inter-regional connectivity and activity), and vascular responses (cerebral blood flow). A secondary aim is to link these neural changes to real-world outcomes, including cognition, learning, and social functioning across adult populations (Figure 1). This synthesis is clinically and socially relevant, as it may clarify whether and how music can serve as a tool to support brain health and well-being, for instance, by informing mental health interventions, optimizing learning, or guiding therapeutic approaches for aging populations at risk of cognitive decline.

Of not the study has some strengths worth mention. The review will follow international systematic review guidelines to ensure rigor and transparency. Multiple major scientific databases will be searched, supplemented by reference screening of prior reviews. Eligible studies will include randomized and non-randomized trials as well as observational designs. Study quality will be appraised using established tools. Anticipated challenges include methodological heterogeneity, variable research quality, and the complexity of cultural, developmental, and music-type influences, which may constrain the ability to generalize findings or conduct meta-analysis.

Despite these limitations, the review will provide an integrated overview of music’s effects on adult brain and behavior, identify key knowledge gaps, and inform future research directions. Findings may support the development of evidence-based interventions to enhance cognition, strengthen emotional resilience, and mitigate age-related decline, thereby advancing the application of music as a therapeutic and preventive strategy for brain health.

## Supporting Information

S1 File. PRISMA-P (Preferred Reporting Items for Systematic Review and Meta-Analysis Protocols) 2015 checklist: Recommended items to address in a systematic review protocol.

## Abbreviation

CBF: Cerebral Blood Flow
CENTRAL: Cochrane Central Register of Controlled Trials
DTI: Diffusion Tensor Imaging
EEG: Electroencephalography
EMBASE: Excerpta Medica Database
GM: Gray Matter
MEG: Magnetoencephalography
MRI: Magnetic Resonance Imaging
NHLBI: National Heart, Lung, and Blood Institute
PRISMA: Preferred Reporting Items for Systematic Reviews and Meta-Analyses
PRISMA-P: Preferred Reporting Items for Systematic Review and Meta-Analysis Protocols
PROSPERO: International Prospective Register of Systematic Reviews
ROBINS-E: Risk of Bias in Non-randomized Studies – of Exposures
ROBINS-I: Risk of Bias in Non-randomized Studies – of Interventions
RoB 2.0: Risk of Bias 2.0
STROBE: Strengthening the Reporting of Observational Studies in Epidemiology
SWiM: Synthesis Without Meta-analysis
WM: White Matter

## Authors Contribution

Conceptualization: I.O.D.J and C.G.; Project administration: I.O.D.J and C.G.; Resources: I.O.D.J., D.S.R., C.G.; Methodology: I.O.D.J and C.G.; Investigation: I.O.D.J and C.G.; Software: I.O.D.J., D.S.R., C.G.; Writing original draft: I.O.D.J and C.G.; Writing reviews and editing: I.O.D.J., D.S.R., C.G.; Correspondence: I.O.D.J and C.G.; Supervision: C.G.

## Data Availability

No new data were generated or collected for the purposes of this study. This protocol relies entirely on information extracted from previously published research and secondary sources. All relevant details pertaining to the study design, methodology, eligibility criteria, data extraction procedures, quality assessment tools, and planned analyses are fully described within the main manuscript. Additionally, supplementary information files provide further clarification and transparency regarding the systematic review process, ensuring that all procedural and methodological aspects are accessible to readers. This approach ensures reproducibility, allows for independent verification, and maintains adherence to standards for rigorous and transparent research reporting.

## Ethical Consideration

This study is a systematic review protocol that relies exclusively on previously published studies and secondary data sources. No new data will be collected from human participants or animals, and no interventions or experimental procedures will be conducted. As such, the review does not involve direct interaction with participants, nor does it include the collection of identifiable personal information. Consequently, ethical approval from an institutional review board or ethics committee was not required. The study will, however, adhere to established ethical standards for research conduct, including transparent reporting, accurate citation of original sources, and responsible synthesis of evidence. All included studies will be appropriately referenced to respect intellectual property and the integrity of the original research.

## Acknowledgements

The author would like to thank Ms. Mary White of the F.D. Bluford Library, North Carolina Agricultural and Technical State University, Greensboro, NC, United States, for her assistance with developing the search terms and strategies.

## Funding

This work did not receive any specific funding from public, commercial, or non-profit organizations.

## Consent for Publication

Not applicable.

## Competing Interests

The authors declare that they have no competing interests.

## Notes

### Competing Interest Statement

The authors have declared no competing interest.

### Funding Statement

The author(s) received no specific funding for this work.

## References

1. Gordon CL, Cobb PR, Balasubramaniam R: Recruitment of the motor system during music listening: An ALE meta-analysis of fMRI data. PloS one 2018, 13(11):e0207213.

2. Lee H, Noppeney U: Long-term music training tunes how the brain temporally binds signals from multiple senses. Proceedings of the National Academy of Sciences 2011, 108(51):E1441–E1450.

3. Trost W, Trevor C, Fernandez N, Steiner F, Frühholz S: Live music stimulates the affective brain and emotionally entrains listeners in real time. Proceedings of the National Academy of Sciences 2024, 121(10):e2316306121.

4. Mongelli V, Dehaene S, Vinckier F, Peretz I, Bartolomeo P, Cohen L: Music and words in the visual cortex: The impact of musical expertise. Cortex 2017, 86:260–274.

5. Møller C, Garza-Villarreal EA, Hansen NC, Højlund A, Bærentsen KB, Chakravarty MM, Vuust P: Audiovisual structural connectivity in musicians and non-musicians: a cortical thickness and diffusion tensor imaging study. Scientific reports 2021, 11(1):4324.

6. Stegemöller EL: Exploring a Neuroplasticity Model of Music Therapy. Journal of Music Therapy 2014, 51(3):211–227.

7. Schlaug G: Musicians and music making as a model for the study of brain plasticity. Progress in brain research 2015, 217:37–55.

8. Chatterjee D, Hegde S, Thaut M: Neural plasticity: The substratum of music-based interventions in neurorehabilitation. NeuroRehabilitation 2021, 48(2):155–166.

9. Münte TF, Altenmüller E, Jäncke L: The musician’s brain as a model of neuroplasticity. Nature Reviews Neuroscience 2002, 3(6):473–478.

10. Musliu A, Berisha B, Latifi D: The impact of music in memory. European Journal of Social Science Education and Research 2017, 4(4):222–227.

11. Snyder B: Music and memory: An introduction: MIT press; 2000.

12. Fernandez NB, Trost WJ, Vuilleumier P: Brain networks mediating the influence of background music on selective attention. Social cognitive and affective neuroscience 2019, 14(12):1441–1452.

13. Jolij J, Meurs M: Music alters visual perception. PloS one 2011, 6(4):e18861.

14. Wan CY, Schlaug G: Music making as a tool for promoting brain plasticity across the life span. The Neuroscientist 2010, 16(5):566–577.

15. Reybrouck M, Brattico E: Neuroplasticity beyond sounds: neural adaptations following long-term musical aesthetic experiences. Brain Sciences 2015, 5(1):69–91.

16. Reybrouck M, Vuust P, Brattico E: Music and brain plasticity: how sounds trigger neurogenerative adaptations. Neuroplasticity Insights of Neural Reorganization 2018, 85.

17. Dalla Bella S: Music and brain plasticity. The Oxford handbook of music psychology 2016:325–342.

18. Olszewska AM, Gaca M, Herman AM, Jednoróg K, Marchewka A: How musical training shapes the adult brain: Predispositions and neuroplasticity. Frontiers in neuroscience 2021, 15:630829.

19. Jaschke AC, Honing H, Scherder EJA: Exposure to a musically-enriched environment; Its relationship with executive functions, short-term memory and verbal IQ in primary school children. PLOS ONE 2018, 13(11):e0207265.

20. Bowmer A, Mason K, Knight J, Welch G: Investigating the Impact of a Musical Intervention on Preschool Children’s Executive Function. Frontiers in Psychology 2018, Volume 9 - 2018.

21. Mendes CG, de Paula JJ, Miranda DM: Effects of Background Music on Attentional Networks of Children With and Without Attention Deficit/Hyperactivity Disorder: Case Control Experimental Study. Interactive Journal of Medical Research 2024, 13(1):e53869.

22. Habibi A, Damasio A, Ilari B, Veiga R, Joshi AA, Leahy RM, Haldar JP, Varadarajan D, Bhushan C, Damasio H: Childhood music training induces change in micro and macroscopic brain structure: results from a longitudinal study. Cerebral Cortex 2018, 28(12):4336–4347.

23. Särkämö T, Ripollés P, Vepsäläinen H, Autti T, Silvennoinen HM, Salli E, Laitinen S, Forsblom A, Soinila S, Rodríguez-Fornells A: Structural Changes Induced by Daily Music Listening in the Recovering Brain after Middle Cerebral Artery Stroke: A Voxel-Based Morphometry Study. Frontiers in Human Neuroscience 2014, Volume 8 - 2014.

24. Sa de Almeida J, Baud O, Fau S, Barcos-Munoz F, Courvoisier S, Lordier L, Lazeyras F, Hüppi PS: Music impacts brain cortical microstructural maturation in very preterm infants: A longitudinal diffusion MR imaging study. Developmental Cognitive Neuroscience 2023, 61:101254.

25. Haslbeck FB, Jakab A, Held U, Bassler D, Bucher H-U, Hagmann C: Creative music therapy to promote brain function and brain structure in preterm infants: A randomized controlled pilot study. NeuroImage: Clinical 2020, 25:102171.

26. Sihvonen AJ, Siponkoski S-T, Martínez-Molina N, Laitinen S, Holma M, Ahlfors M, Kuusela L, Pekkola J, Koskinen S, Särkämö T: Neurological Music Therapy Rebuilds Structural Connectome after Traumatic Brain Injury: Secondary Analysis from a Randomized Controlled Trial. Journal of Clinical Medicine 2022, 11(8):2184.

27. Blood AJ, Zatorre RJ: Intensely pleasurable responses to music correlate with activity in brain regions implicated in reward and emotion. Proceedings of the National Academy of Sciences 2001, 98(20):11818–11823.

28. Faßhauer C, Frese A, Evers S: Musical ability is associated with enhanced auditory and visual cognitive processing. BMC neuroscience 2015, 16(1):59.

29. Meyer M, Elmer S, Ringli M, Oechslin MS, Baumann S, Jancke L: Long-term exposure to music enhances the sensitivity of the auditory system in children. European journal of neuroscience 2011, 34(5):755–765.

30. Xu J, Yu L, Cai R, Zhang J, Sun X: Early auditory enrichment with music enhances auditory discrimination learning and alters NR2B protein expression in rat auditory cortex. Behavioural brain research 2009, 196(1):49–54.

31. Humphrey T: The effect of music ear training upon the auditory discrimination abilities of trainable mentally retarded adolescents. Journal of Music Therapy 1980, 17(2):70–74.

32. Rochette F, Moussard A, Bigand E: Music lessons improve auditory perceptual and cognitive performance in deaf children. Frontiers in human neuroscience 2014, 8:488.

33. Karpati FJ, Giacosa C, Foster NEV, Penhune VB, Hyde KL: Sensorimotor integration is enhanced in dancers and musicians. Experimental Brain Research 2016, 234(3):893–903.

34. Hsu H-Y, Lin C-W, Lin Y-C, Wu P-T, Kato H, Su F-C, Kuo L-C: Effects of vibrotactile-enhanced music-based intervention on sensorimotor control capacity in the hand of an aging brain: a pilot feasibility randomized crossover trial. BMC geriatrics 2021, 21(1):660.

35. Carrer LRJ, Pompéia S, Miranda MC: Sensorimotor synchronization with music and metronome in school-aged children. Psychology of Music 2023, 51(2):523–540.

36. Rodriguez-Gomez DA, Talero-Gutiérrez C: Effects of music training in executive function performance in children: A systematic review. Frontiers in Psychology 2022, Volume 13 - 2022.

37. Luo C, Guo Z-w, Lai Y-x, Liao W, Liu Q, Kendrick KM, Yao D-z, Li H: Musical training induces functional plasticity in perceptual and motor networks: insights from resting-state FMRI. PLoS one 2012, 7(5):e36568.

38. Wu J, Zhang J, Ding X, Li R, Zhou C: The effects of music on brain functional networks: a network analysis. Neuroscience 2013, 250:49–59.

39. Alluri V, Toiviainen P, Burunat I, Kliuchko M, Vuust P, Brattico E: Connectivity patterns during music listening: Evidence for action-based processing in musicians. Human brain mapping 2017, 38(6):2955–2970.

40. Meyer M, Elmer S, Ringli M, Oechslin MS, Baumann S, Jancke L: Long-term exposure to music enhances the sensitivity of the auditory system in children. European Journal of Neuroscience 2011, 34(5):755–765.

41. Humphrey T: The Effect of Music Ear Training Upon the Auditory Discrimination Abilities of Trainable Mentally Retarded Adolescents*. Journal of Music Therapy 1980, 17(2):70–74.

42. Chan AS, Ho Y-C, Cheung M-C: Music training improves verbal memory. Nature 1998, 396(6707):128-128.

43. Franklin MS, Sledge Moore K, Yip C-Y, Jonides J, Rattray K, Moher J: The effects of musical training on verbal memory. Psychology of Music 2008, 36(3):353–365.

44. Taylor AC, Dewhurst SA: Investigating the influence of music training on verbal memory. Psychology of Music 2017, 45(6):814–820.

45. Kasuya-Ueba Y, Zhao S, Toichi M: The Effect of Music Intervention on Attention in Children: Experimental Evidence. Frontiers in Neuroscience 2020, Volume 14 - 2020.

46. Pasiali V, LaGasse AB, Penn SL: The Effect of Musical Attention Control Training (MACT) on Attention Skills of Adolescents with Neurodevelopmental Delays: A Pilot Study. Journal of Music Therapy 2014, 51(4):333–354.

47. Moore KS: A Systematic Review on the Neural Effects of Music on Emotion Regulation: Implications for Music Therapy Practice. Journal of Music Therapy 2013, 50(3):198–242.

48. Cook T, Roy ARK, Welker KM: Music as an emotion regulation strategy: An examination of genres of music and their roles in emotion regulation. Psychology of Music 2019, 47(1):144–154.

49. Sharda M, Tuerk C, Chowdhury R, Jamey K, Foster N, Custo-Blanch M, Tan M, Nadig A, Hyde K: Music improves social communication and auditory–motor connectivity in children with autism. Translational psychiatry 2018, 8(1):231.

50. Boster JB, Spitzley AM, Castle TW, Jewell AR, Corso CL, McCarthy JW: Music improves social and participation outcomes for individuals with communication disorders: A systematic review. Journal of Music Therapy 2021, 58(1):12–42.

51. Bernatzky G, Bernatzky P, Hesse H-P, Staffen W, Ladurner G: Stimulating music increases motor coordination in patients afflicted with Morbus Parkinson. Neuroscience Letters 2004, 361(1):4–8.

52. Arnaud.Cabanac, Perlovsky L, Bonniot-Cabanac M-C, Cabanac M: Music and academic performance. Behavioural Brain Research 2013, 256:257–260.

53. Li H-C, Wang H-H, Chou F-H, Chen K-M: The effect of music therapy on cognitive functioning among older adults: A systematic review and meta-analysis. Journal of the American Medical Directors Association 2015, 16(1):71–77.

54. Román-Caballero R, Arnedo M, Triviño M, Lupiáñez J: Musical practice as an enhancer of cognitive function in healthy aging - A systematic review and meta-analysis. PLOS ONE 2018, 13(11):e0207957.

55. Hyde KL, Lerch J, Norton A, Forgeard M, Winner E, Evans AC, Schlaug G: The Effects of Musical Training on Structural Brain Development. Annals of the New York Academy of Sciences 2009, 1169(1):182–186.

56. Fischer CE, Churchill N, Leggieri M, Vuong V, Tau M, Fornazzari LR, Thaut MH, Schweizer TA: Long-known music exposure effects on brain imaging and cognition in early-stage cognitive decline: A pilot study. Journal of Alzheimer’s Disease 2021, 84(2):819–833.

57. Gaser C, Schlaug G: Brain structures differ between musicians and non-musicians. Journal of neuroscience 2003, 23(27):9240–9245.

58. Zhang L, Peng W, Chen J, Hu L: Electrophysiological evidences demonstrating differences in brain functions between nonmusicians and musicians. Scientific Reports 2015, 5(1):13796.

59. Schmithorst VJ, Wilke M: Differences in white matter architecture between musicians and non-musicians: a diffusion tensor imaging study. Neuroscience letters 2002, 321(1-2):57–60.

60. Zuk J, Benjamin C, Kenyon A, Gaab N: Behavioral and neural correlates of executive functioning in musicians and non-musicians. PloS one 2014, 9(6):e99868.

61. Schulze K, Müller K, Koelsch S: Neural correlates of strategy use during auditory working memory in musicians and non-musicians. European Journal of Neuroscience 2011, 33(1):189–196.

62. Partanen E, Kujala T, Tervaniemi M, Huotilainen M: Prenatal music exposure induces long-term neural effects. PloS one 2013, 8(10):e78946.

63. Mahmood D, Nisar H, Yap VV, Tsai C-Y: The effect of music listening on EEG functional connectivity of brain: A short-duration and long-duration study. Mathematics 2022, 10(3):349.

64. Raglio A, Galandra C, Sibilla L, Esposito F, Gaeta F, Di Salle F, Moro L, Carne I, Bastianello S, Baldi M: Effects of active music therapy on the normal brain: fMRI based evidence. Brain imaging and behavior 2016, 10(1):182–186.

65. Steinhoff N, Heine AM, Vogl J, Weiss K, Aschraf A, Hajek P, Schnider P, Tucek G: A pilot study into the effects of music therapy on different areas of the brain of individuals with unresponsive wakefulness syndrome. Frontiers in Neuroscience 2015, Volume 9 - 2015.

66. Lanzilotti C, Dumas R, Grassi M, Schön D: Prolonged exposure to highly rhythmic music affects brain dynamics and perception. Neuropsychologia 2019, 129:191–199.

67. Xiao X, Chen W, Zhang X: The effect and mechanisms of music therapy on the autonomic nervous system and brain networks of patients of minimal conscious states: a randomized controlled trial. Frontiers in neuroscience 2023, 17:1182181.

68. Moore E, Schaefer RS, Bastin ME, Roberts N, Overy K: Diffusion tensor MRI tractography reveals increased fractional anisotropy (FA) in arcuate fasciculus following music-cued motor training. Brain and cognition 2017, 116:40–46.

69. Fachner J, Gold C, Erkkilä J: Music therapy modulates fronto-temporal activity in rest-EEG in depressed clients. Brain topography 2013, 26(2):338–354.

70. Bhattacharya J, Lee E-J: Modulation of EEG theta band signal complexity by music therapy. International Journal of Bifurcation and Chaos 2016, 26(01):1650001.

71. Nawaz R, Nisar H, Voon YV: The effect of music on human brain; Frequency domain and time series analysis using electroencephalogram. Ieee Access 2018, 6:45191–45205.

72. Hauck M, Metzner S, Rohlffs F, Lorenz J, Engel AK: The influence of music and music therapy on pain-induced neuronal oscillations measured by magnetencephalography. Pain 2013, 154(4):539–547.

73. Buard I, Dewispelaere WB, Thaut M, Kluger BM: Preliminary neurophysiological evidence of altered cortical activity and connectivity with neurologic music therapy in Parkinson’s disease. Frontiers in neuroscience 2019, 13:105.

74. Fujioka T, Chen JL, Black SE, Chen JJ, Honjo K, Dawson DR, Ross B: Beta-and gamma-band neuromagnetic oscillations in chronic stroke rehabilitation using music-supported therapy and manual training. Annals of the New York Academy of Sciences 2025.

75. Rajakumar KD, Mohan J: A systematic review on effect of music intervention on cognitive impairment using EEG, fMRI, and cognitive assessment modalities. Results in Engineering 2024, 22:102224.

76. Feng K, Shen C-Y, Ma X-Y, Chen G-F, Zhang M-L, Xu B, Liu X-M, Sun J-J, Zhang X-Q, Liu P-Z: Effects of music therapy on major depressive disorder: A study of prefrontal hemodynamic functions using fNIRS. Psychiatry research 2019, 275:86–93.

77. Gutiérrez Santamaría S: Microstructural changes related to dementia and music therapy in memory impaired patients measured with diffusion MRI: first insights of the ALMUTH trial. 2022.

78. Siponkoski S-T, Martinez-Molina N, Kuusela L, Laitinen S, Holma M, Ahlfors M, Jordan-Kilkki P, Ala-Kauhaluoma K, Melkas S, Pekkola J: Music therapy enhances executive functions and prefrontal structural neuroplasticity after traumatic brain injury: evidence from a randomized controlled trial. Journal of neurotrauma 2020, 37(4):618–634.

79. Braz CH, Gonçalves LF, Paiva KM, Haas P, Patatt FSA: Implications of musical practice in central auditory processing: a systematic review. Brazilian Journal of Otorhinolaryngology 2021, 87(2):217–226.

80. Kaiser AP, Berntsen D: The cognitive characteristics of music-evoked autobiographical memories: Evidence from a systematic review of clinical investigations. Wiley Interdisciplinary Reviews: Cognitive Science 2023, 14(3):e1627.

81. Talamini F, Altoè G, Carretti B, Grassi M: Musicians have better memory than nonmusicians: A meta-analysis. PloS one 2017, 12(10):e0186773.

82. Martin-Moratinos M, Bella-Fernandez M, Blasco-Fontecilla H: Effects of music on attention-deficit/hyperactivity disorder (ADHD) and potential application in serious video games: systematic review. Journal of medical Internet research 2023, 25:e37742.

83. Chee ZJ, Chang CYM, Cheong JY, Malek FHBA, Hussain S, de Vries M, Bellato A: The effects of music and auditory stimulation on autonomic arousal, cognition and attention: a systematic review. International Journal of Psychophysiology 2024, 199:112328.

84. Liu Q, Li W, Yin Y, Zhao Z, Yang Y, Zhao Y, Tan Y, Yu J: The effect of music therapy on language recovery in patients with aphasia after stroke: a systematic review and meta-analysis. Neurological Sciences 2022, 43(2):863–872.

85. Yang Y, Fang Y-Y, Gao J, Geng G-L: Effects of five-element music on language recovery in patients with poststroke aphasia: a systematic review and meta-analysis. The Journal of Alternative and Complementary Medicine 2019, 25(10):993–1004.

86. Uhlig S, Jaschke A, Scherder E: Effects of music on emotion regulation: A systematic literature review. 2013.

87. Kawasaki A, Hayashi N: Playing a musical instrument increases blood flow in the middle cerebral artery. PLOS ONE 2022, 17(6):e0269679.

88. Cavieres R, Landerretche J, Jara JL, Chacón M: Analysis of cerebral blood flow entropy while listening to music with emotional content. Physiological Measurement 2021, 42(5):055002.

89. Ayaz A, Rahimi A, Buwadi L, Wang Y-B, Zou L, Heath M: Rocking the cerebral blood flow: the influence of music listening and aerobic exercise on cortical hemodynamics and post-intervention executive function. Experimental Brain Research 2025, 243(4):102.

90. Paulson OB, Hasselbalch SG, Rostrup E, Knudsen GM, Pelligrino D: Cerebral Blood Flow Response to Functional Activation. Journal of Cerebral Blood Flow & Metabolism 2010, 30(1):2–14.

91. Lauritzen M: Relationship of spikes, synaptic activity, and local changes of cerebral blood flow. Journal of Cerebral Blood Flow & Metabolism 2001, 21(12):1367–1383.

92. Nakamura S, Sadato N, Oohashi T, Nishina E, Fuwamoto Y, Yonekura Y: Analysis of music–brain interaction with simultaneous measurement of regional cerebral blood flow and electroencephalogram beta rhythm in human subjects. Neuroscience letters 1999, 275(3):222–226.

93. Blood AJ, Zatorre RJ, Bermudez P, Evans AC: Emotional responses to pleasant and unpleasant music correlate with activity in paralimbic brain regions. Nature neuroscience 1999, 2(4):382–387.

94. Altenmüller E, Marco-Pallares J, Münte T, Schneider S: Neural reorganization underlies improvement in stroke-induced motor dysfunction by music-supported therapy. Annals of the New York Academy of Sciences 2009, 1169(1):395–405.

95. Page MJ, McKenzie JE, Bossuyt PM, Boutron I, Hoffmann TC, Mulrow CD, Shamseer L, Tetzlaff JM, Akl EA, Brennan SE: The PRISMA 2020 statement: an updated guideline for reporting systematic reviews. bmj 2021, 372.

96. Campbell M, McKenzie JE, Sowden A, Katikireddi SV, Brennan SE, Ellis S, Hartmann-Boyce J, Ryan R, Shepperd S, Thomas J: Synthesis without meta-analysis (SWiM) in systematic reviews: reporting guideline. bmj 2020, 368.

97. Moher D, Shamseer L, Clarke M, Ghersi D, Liberati A, Petticrew M, Shekelle P, Stewart LA, Group P-P: Preferred reporting items for systematic review and meta-analysis protocols (PRISMA-P) 2015 statement. Systematic reviews 2015, 4(1):1.

98. Shamseer L, Moher D, Clarke M, Ghersi D, Liberati A, Petticrew M, Shekelle P, Stewart LA: Preferred reporting items for systematic review and meta-analysis protocols (PRISMA-P) 2015: elaboration and explanation. Bmj 2015, 349.

99. Minozzi S, Cinquini M, Gianola S, Gonzalez-Lorenzo M, Banzi R: The revised Cochrane risk of bias tool for randomized trials (RoB 2) showed low interrater reliability and challenges in its application. Journal of clinical epidemiology 2020, 126:37–44.

100. Jüni P, Loke Y, Pigott T, Ramsay C, Regidor D, Rothstein H, Sandhu L, Santaguida P, Schünemann H, Shea B: Risk of bias in non-randomized studies of interventions (ROBINS-I): detailed guidance. Br Med J 2016, 355:i4919.

101. Higgins JP, Morgan RL, Rooney AA, Taylor KW, Thayer KA, Silva RA, Lemeris C, Akl EA, Bateson TF, Berkman ND: A tool to assess risk of bias in non-randomized follow-up studies of exposure effects (ROBINS-E). Environment international 2024, 186:108602.

102. Cevallos M, Egger M: STROBE (STrengthening the Reporting of OBservational studies in Epidemiology). Guidelines for reporting health research: a user’s manual 2014:169–179.

103. Page MJ, McKenzie JE, Bossuyt PM, Boutron I, Hoffmann TC, Mulrow CD, Shamseer L, Tetzlaff JM, Akl EA, Brennan SE et al: The PRISMA 2020 statement: an updated guideline for reporting systematic reviews. BMJ 2021, 372:n71.

104. Shao S-C, Kuo L-T, Huang Y-T, Lai P-C, Chi C-C: Using Grading of Recommendations Assessment, Development, and Evaluation (GRADE) to rate the certainty of evidence of study outcomes from systematic reviews: A quick tutorial. Dermatologica Sinica 2023, 41(1):3–7.

105. Toader C, Tataru CP, Florian I-A, Covache-Busuioc R-A, Bratu B-G, Glavan LA, Bordeianu A, Dumitrascu D-I, Ciurea AV: Cognitive crescendo: how music shapes the brain’s structure and function. Brain Sciences 2023, 13(10):1390.

106. Creutzfeldt O, Ojemann G: Neuronal activity in the human lateral temporal lobe: III. Activity changes during music. Experimental Brain Research 1989, 77(3):490–498.

107. Freitas C, Manzato E, Burini A, Taylor MJ, Lerch JP, Anagnostou E: Neural correlates of familiarity in music listening: A systematic review and a neuroimaging meta-analysis. Frontiers in neuroscience 2018, 12:686.

108. Zhang Y, Cai J, An L, Hui F, Ren T, Ma H, Zhao Q: Does music therapy enhance behavioral and cognitive function in elderly dementia patients? A systematic review and meta-analysis. Ageing research reviews 2017, 35:1–11.

109. Ueda T, Suzukamo Y, Sato M, Izumi S-I: Effects of music therapy on behavioral and psychological symptoms of dementia: a systematic review and meta-analysis. Ageing research reviews 2013, 12(2):628–641.

